# A Phase 2 random, double-blind, placebo-controlled study of the safety and immunogenicity of a recombinant G protein-based respiratory syncytial virus vaccine in healthy older adults

**DOI:** 10.1101/2023.10.26.23297584

**Authors:** Lunan Zhang, Gan Zhao, Xin Cheng, Shuo Wang, Jiarong Wang, Xuefen Huai, Yu Xia, Yanling Xiao, Sulin Ren, Shijie Zhang, Bin Wang

## Abstract

**Background:** Respiratory syncytial virus (RSV) is an important cause of disease in older adults. Vaccines against RSV infections and respiratory diseases are in large market demand. Although there are currently two licensed RSV-based pre-F antigen vaccines available for older adults, no G antigen-based RSV vaccine is authorized. This phase 2 study aimed to evaluate the safety and immunogenicity of a recombinant G protein-based RSV vaccine in this population.

**Methods:** A phase-2 randomized, double-blind, placebo-controlled, dose-ranging study was conducted to evaluate the safety, tolerability and immunogenicity of the BARS13 (rRSV G protein with CsA) when administered by an intramuscular (IM) injection to healthy participants 60 to 80 years old. A total of 125 eligible participants were randomized in a 3:1 ratio (vaccine versus placebo) for Cohorts 1 and 2 and randomized in a 2:1 ratio for Cohort 3 to receive one of the three treatment regimens or placebo.

**Results:** The average age was 65.3, and 50.4% (63/125) were men. Until the interim analysis (4 weeks following the last vaccination), no treatment-related SAE occurred. TEAEs did not increase with vaccination dosage or frequency. All adverse effects were mild or moderate, not severe or life-threatening. BARS13 vaccination increased IgG anti-RSV antibody levels in all cohorts, but higher doses and frequency boosted immune responses significantly. The high-dose thrice-administered recipients had serum-specific IgG antibody GMC of 881.0 IU/mL (95% CI: 794.5-1473.4) before the first dose (Week 0), 1116.3 IU/mL (95% CI:990.7-1772.5) 4 weeks after the first dose (Week 4), 1309.4IU/mL (95% CI: 1162.8-2041.5) 4 weeks after the second dose (Week 8), and1359.6 IU/mL (95% CI: 1197.9-2525.7) 4 weeks after the third dose (Week 12). For the low-dose twice-administered recipients, 84% responded at 4 weeks after the first immunization (Week 4) and 83.3% at 4 weeks after the second (Week 8). The high-dose twice-administered recipients had 95.5% response at 4 weeks after the first immunization (Week 4) and 72.2% at 4 weeks after the second (Week 8). At Week 4, 85.7% of high-dose thrice-administered recipients responded, 85.2% at Week 8, and 79.2% at Week 12.

**Conclusions:** The study demonstrates the safety and tolerability of BARS13 across different dose groups. Adverse reactions were not significantly different among participants receiving varying doses of BARS13. Levels of anti-G antibodies exhibited a dose- and frequency-dependent responses in the older population. The continuous upward trend in antibody concentration up to the interim analysis is promising for the effectiveness of BARS13.

## 1. BACKGROUND

A growing number of complications have been observed in the older adult population as a consequence of the infection caused by respiratory syncytial virus (RSV). Based on a number of studies conducted in various regions of China utilizing health care databases and viral surveillance data, it has been observed that individuals aged 60 years and older who contract RSV infections account for the second highest proportion (25.2%) of the total annual RSV infections. Furthermore, this age group exhibits a higher likelihood of hospitalization, increased rates of admission to intensive care units, longer durations of hospital stay, and elevated mortality rates when compared to children infected with RSV [1–3]. In the United States, RSV was also estimated to be the causative agent in up to 12% of acute respiratory illnesses in older adults who were not selected for comorbidities. The prevalence of RSV varied depending on the clinical context and the year of study [4]. Treatments for RSV in older adults are currently limited to supportive care, including oxygen therapy and more advanced breathing support with continuous positive airway pressure (CPAP) or nasal high-flow oxygen, as no antiviral therapy is approved for the condition. Vaccination remains a high priority.

The development of a formalin-inactivated RSV (FI-RSV) vaccine for newborns and young age groups was problematic in the early 1960s due to the observation of vaccine-enhanced disease (VED) after administration of the FI-RSV vaccine [5–7]. Different vaccine strategies have been considered and investigated for preventing severe RSV infections since then. Until recently, structural biological studies demonstrated stabilized pre-F protein subunit vaccines to be a promising approach for RSV vaccination [8]. These subunit vaccines aim to target the pre-fusion conformation of the RSV fusion (F) protein, which has been shown to induce a stronger neutralizing antibody response compared to the post-fusion form, both in preclinical and clinical settings [9, 10]. This breakthrough in vaccine design has revitalized efforts towards developing an effective and safe RSV vaccine. Two pre-F protein subunit vaccines were successfully tested in phase III clinical trials in older populations, with approximate 66-85% and 71-94% protections against RSV-related lower respiratory disease [11, 12]. Currently, the FDA has approved two RSV subunit vaccines for older adults for marketing. These vaccines have been shown to be effective in preventing RSV-related respiratory illnesses in older populations, offering great hope for combating this virus. The FDA’s approval of these vaccines for marketing is a significant step forward in addressing the need for an effective and safe RSV vaccine.

With ongoing research and advancements in vaccine design, it is hopeful that similar success can be achieved in developing preventative measures for other vulnerable populations, such as infants and young children. However, it is important to note that the effectiveness of the RSV vaccines in older populations may not necessarily translate to the same level of success in infants and young children. This is due to differences in immune response and susceptibility to the virus between age groups [13]. Therefore, further research and clinical trials specifically targeting these vulnerable populations are necessary to determine the efficacy of RSV vaccines in preventing respiratory illnesses in infants and young children. Although the pre-F-based vaccine design has been successful, other non-fusion antigens are still considerable for developing as RSV vaccine candidate. For example, the glycoprotein G protein of RSV functions as an attachment protein during RSV infection by interacting with the receptors of target cells. The attachment of the glycoprotein G protein to the receptor allows the virus to enter the host’s cells and initiate infection [14]. By targeting the RSV G protein in a subunit vaccine, researchers aim to induce an immune response that can neutralize the virus before it enters the cells, preventing infection. This approach has shown promising results in preclinical studies, underscoring the potential of the G protein as a feasible contender for the advancement of a subunit respiratory syncytial virus (RSV) vaccine [15–17]. In addition, the bacterially produced un-glycosylated G protein could protect against homologous and heterologous RSV challenges in mice, suggesting its potential as a broad-spectrum vaccine candidate [18]. Furthermore, the use of the un-glycosylated G protein in a subunit vaccine may offer advantages such as improved stability and cost effects, making it a promising option for large-scale production and distribution.

We recently conducted the first-in-human phase 1 study that evaluated un-glycosylated G protein with a CsA as an immunomodulator as the BARS13RSV vaccine in healthy adults with different dose levels. The results showed no serious adverse events in 18 to 45-year-old volunteers. The vaccine induced a strong humoral immune response, with increased levels of RSV-specific antibodies [19]. The recombinant G protein is a promising candidate for developing a subunit RSV vaccine, and further testing is needed in the elderly population. A phase 2 study is currently underway to determine if BARS13 would result in a stronger immune response in older adults.

## 2. METHODS

### 2.1 Study Design

The goal of this phase 2 randomized, double-blind, placebo-controlled, dose-ranging study (NCT04681833) is to find out if BARS13 (rRSV G protein with CsA) is safe and well tolerated when injected into healthy people between the ages of 60 and 80. The study follows a low-dose duplicate (10 µg BARS13 administered on days 1 and 29), high-dose duplicate (20 µg BARS13 administered on days 1 and 29), or high-dose triplicate (20 µg BARS13 administered on days 1, 29, and 57) vaccination schedule. The study was conducted at two sites in Australia. The results presented here are relevant to the interim analysis, which occurred 4 weeks post last dose (FU3) for each cohort.

The clinical protocol was approved by the Bellberry Human Research Ethics Committee. The conduct of the study followed the Declaration of Helsinki and the current Good Clinical Practices. All participants provided written informed consent prior to enrollment. Human serum samples were prepared at Mater Pathology in Queensland, Australia, and Agilex Biolabs in Adelaide, Australia, the latter performed the immunological tests in this experiment at the same time.

The primary endpoints focused on safety and tolerability by assessing the incidence and severity of side effects related to the vaccine, both locally and systemically. The secondary endpoint was to measure anti-G protein specific IgG antibody levels using an enzyme-linked immunosorbent assay (ELISA).

As specified in the protocol, an unblinded interim analysis is being carried out by an independent team, and the results will be shared with the unblinded sponsor team to maintain the blindness of the entire study. Staff managing the general aspects of the trial at the sites, the CRO, and the sponsor will remain blinded.

### 2.2 Participants

Healthy elderly male and female subjects between the ages of 60 to 80 years with no history of severe allergies or immunosuppressive therapy were enrolled. All participants provided written informed consent before participation. Key exclusion criteria included known infection with HIV, HBV, or HCV, receipt of any other vaccines within 4 weeks before entering the study or 4 weeks within the last dose of study vaccine, and any previous investigational RSV vaccination. Influenza vaccine and coronavirus SARS-CoV-2 (COVID-19) vaccine should not be administered within a 14-day interval of each dose of study vaccine. Participants were assessed at the screening period, dose administration period, follow-up visits, and final follow-up/end-of-study visit.

### 2.3 The Vaccine

The lyophilized powder of rRSV G protein and CsA diluent was produced by Advaccine Biopharmaceuticals Suzhou Co. Ltd., located in Suzhou, China. The formulation buffer without active components was used as placebo. The rRSV G protein lyophilized powder and sterile CsA diluent solution were mixed together as the active BARS13 vaccine for injection. Details about the study vaccine lots can be found in Supplementary File S1.

### 2.4 Study Procedures

The study recruited and randomly allocated eligible participants in Cohorts 1 and 2 in a 3:1 ratio (BARS13 versus placebo, 30 active, 10 placebo in each cohort) and in Cohort 3 in a 2:1 ratio (30 active, 15 placebo). Participants in Cohort 1 received low-dose BARS13 (10 µg) or placebo on days 1 and 29, in Cohort 2 received high-dose BARS13 (20 µg) or placebo on days 1 and 29, while in Cohort 3 received high-dose BARS13 (20 µg) or placebo on days 1, 29 and 57. Follow-up visits were scheduled for 1 week (days 7 and 36 for all cohorts, while days 64 for Cohort 3 only), 2 weeks (days 15 and 43 for all cohorts, while days 71 for Cohort 3 only) and 4 weeks (days 29 and 57 for all cohorts, while days 85 for Cohorts 3 only) post each vaccination.

As part of the study protocol, an interim analysis was set up to observe the safety, tolerability, and immunogenicity of all cohorts up to 4 weeks after their last immunization.

### 2.5 Safety Assessment

All safety endpoints were analyzed using the safety population. The severity and relationship of adverse events (AEs) to the vaccine regimens were assessed by the investigators based on the US Food and Drug Administration (FDA) standards (FDA 2007, Guidance for Industry: Toxicity Grading Scale for Health Adult and Adolescent Volunteers Enrolled in Preventive Vaccine Clinical Trials). Treatment-emergent AEs (TEAEs) were defined as AEs that occurred or worsened from the first administration of the study vaccine up to 28 days after the last dose. If a determination couldn’t be made as to whether the event was treatment emergent due to missing or incomplete data, the adverse event would be treated as treatment emergent. For maximum severity/causality of TEAE, the maximum severity/causality of each participant across all TEAEs were counted. A treatment-related (Serious) TEAEs was one for which the causality of a study vaccine administration-related ‘Possibly’, ‘Probably’, or ‘Definitely’ was established. The number of participants with the percentage (%) of each treatment group were displayed in the corresponding outputs. All AE summaries were restricted to TEAEs only. Counts and percentage of systemic reactions (such as fatigue, myalgia and malaise) and injection site reactions (such as pain, tenderness and erythema) were summarized by reaction type at each protocol specified timepoint for treatment group under the safety population.

### 2.6 Immunogenicity Assessment

An immunogenicity assessment was conducted to evaluate the immune response elicited by the vaccinations. Serum samples from participants were collected at specified time points and analyzed for the presence of antibodies against the recombinant G protein (IgG against rRSV G protein). Serum samples from Cohorts 1 and 2 were collected on Day 1 (Week 0), Day 29 (Week 4), Day 57 (Week 8), while from Cohort 3 were on Day 1 (Week 0), Day 29 (Week 4), Day 57 (Week 8), and Day 85 (Weeks 12). The humoral immune responses were analyzed for the presence of immunoglobulin G (IgG) antibodies targeting the RSV G protein, as indicated by the data gathered for the interim analysis. The immunogenicity assessment assay was validated and conducted in accordance with a validated method and analysis results were presented as the geometric mean concentration values in each group and the proportion of participants who had an increase in antibody concentration after vaccine immunization. Following the final visit of this trial, additional tests will be performed and analyzed in a consistent manner.

The quantitative enzyme-linked immunosorbent test (ELISA) was used to assess the levels of G protein-specific IgG antibodies. Plates were first coated with rRSV protein G, following by a blocking step. The standard curve was established by serially diluting a standard RSV IgG serum (NIBSC, London, UK, Cat No.:16/284). Human serum samples were diluted (1 in 2000) and placed on the plate for a 1-hour incubation. Following washing, goat anti-human IgG (H+L) peroxidase-labeled anti-protein G IgG antibodies (Invitrogen, Carlsbad, CA, USA, Catalog No.: 31410) were added to the plate for a further 1-hour incubation, followed by additional washes. TMB (Sigma-Aldrich, St. Louis, MO, USA, Catalog No.: T0440) and a stop solution were used to create a colorimetric signal. An ELISA plate reader (SpectraMax VersaMax, Molecular Devices, Sunnyvale, CA, USA) was used to read the signal. The signal produced was directly proportional to the quantity of analyte present and was determined by interpolating from the calibration curve provided on each plate. The concentrations of anti-G protein IgG antibodies in the samples were determined using calibration curves, specifically employing a four parameter logistics (4-PL) curve fitting method with a weighting factor of 1/Y. The data were then exported to GraphPad Prism (GraphPad, San Diego, CA, USA, version 9.3) for further analysis.

### 2.7 Statistical Analysis

There were only descriptive statistics performed for the safety data as the main result, and no statistical hypothesis was developed for this analysis. Immunogenicity data gathered during both the pre-dose and post-dose periods were meticulously cleaned, summarized, and subjected to statistical analysis. Numerical data, collected at predetermined intervals, were consolidated and statistically scrutinized in accordance with their respective cohorts. Given the fact that the test results were taken from unblinded data, it is postulated that the levels of antibodies in those who were administered the placebo would remain constant during the pre-dose and post-dose periods. Hence, based on the distribution of participants within the placebo group, we excluded data from equivalent percentages of people whose antibody levels exhibited insignificant changes before and after vaccination from the analysis. The data underwent analysis and output using GraphPad Prism 9.3 software. Statistical analysis was performed using a paired T-test. All statistical tests conducted in this study were two-sided, and any observed differences with a *p*-value less than 0.05 were deemed statistically significant.

## 3. RESULTS

### 3.1 Participant Demographics

From May 19, 2021, to December 19, 2022, an enrollment screening was conducted on a total of 223 participants. A total of 127 participants were enrolled and randomized, as illustrated in Figure 1. Of these, 125 were designated as allocated cohorts. Of the 125 participants overall, 124 (99.2%) participants received the allocated treatment, and 125 (100.0%) participants completed the FU3 visit, which was 4 weeks after the last dose.

**Figure 1.**
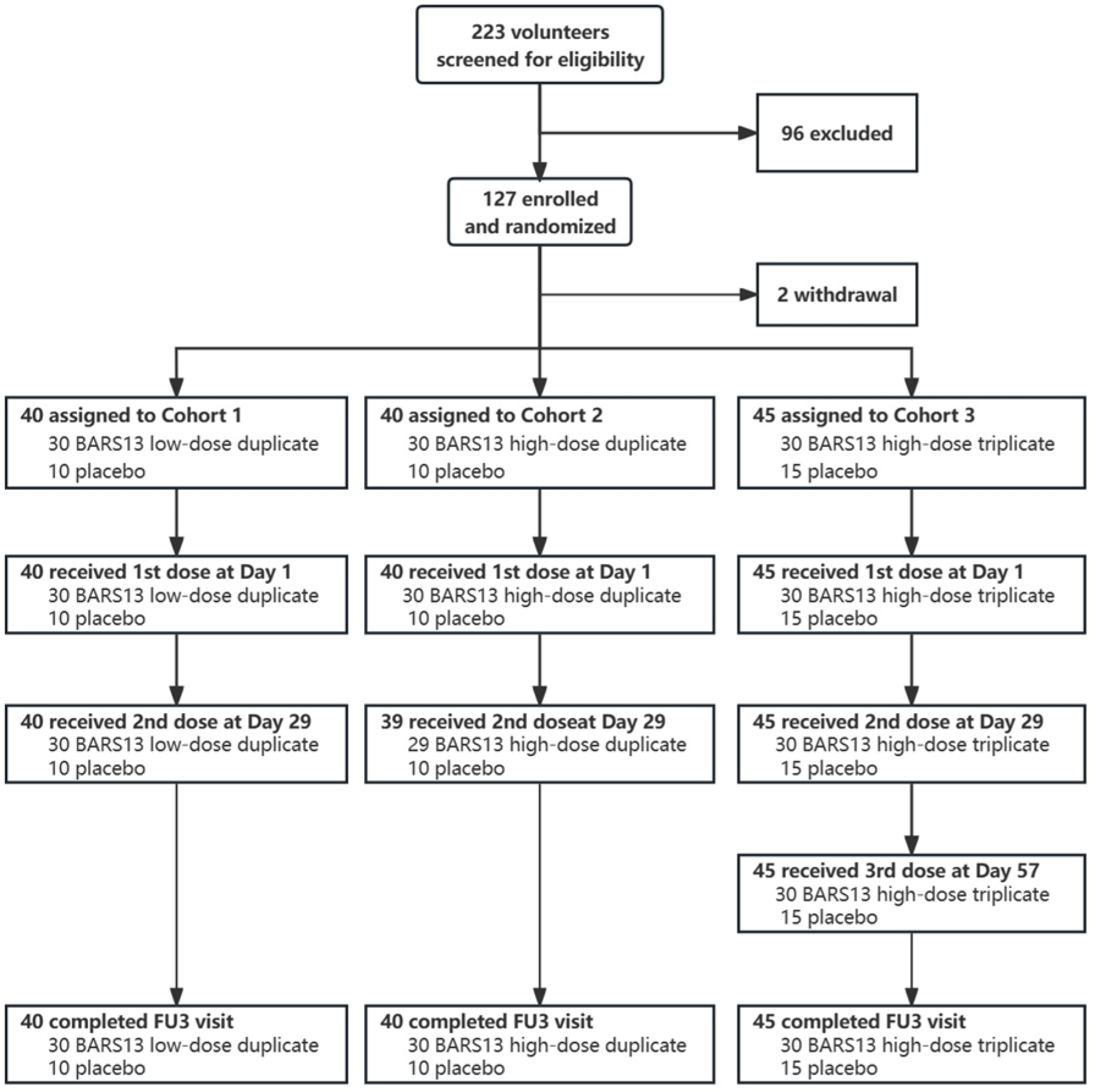
Enrollment and Follow-up of the Participants. The data cutoff timepoint for Cohorts 1 and 2 was on Day 57, but for Cohort 3, represented by FU3, it was on Day 85. A total of 125 participants, ranging in age from 60 to 80, were subjected to a randomization process in order to be assigned to one of two groups. The first group received BARS13 at different dosage levels, while the second group received a placebo. The safety population consisted of all subjects who were administered either the research therapy BARS13 or a placebo. At the point of data cutoff, a total of 125 participants were under observation.

The demographic and baseline characteristics were generally balanced and comparable across all treatment groups (Table 1). Out of all the 125 participants in the safety population, 63 were male (50.4%) and 62 were female (49.6%). The average age was 65.3 years old (ranging from 60 to 77 years old). The mean BMI was 28.4 kg/m^2^ (ranging from 19 to 40 kg/m^2^). 121 participants (96.8%) were white, and 3 participants (2.4%) were Asian.

**Table 1.** Demographics and Baseline Characteristics (Safety Population)

| **Characteristics** | **LDR**<br>**(N=30)** | **HDR**<br>**(N=30)** | **HTR**<br>**(N=30)** | **Pooled**<br>**Placebo**<br>**(N=35)** | **Overall**<br>**(N=125)** |
| --- | --- | --- | --- | --- | --- |
| **Age (years)** | 65.2 (4.87) | 65.7 (4.29) | 64.3 (4.07) | 65.9 (4.63) | 65.3 (4.46) |
| **BMI at screening (kg/m<sup>2</sup>)** | 27.9 (3.68) | 28.7 (4.64) | 29.2 (4.42) | 28.0 (5.46) | 28.4 (4.60) |
| **Gender** |  |  |  |  |  |
| Male | 16 (53.3%) | 12 (40.0%) | 18 (60.0%) | 17 (48.6%) | 63 (50.4%) |
| Female | 14 (46.7%) | 18 (60.0%) | 12 (40.0%) | 18 (51.4%) | 62 (49.6%) |
| **Race** |  |  |  |  |  |
| Asian | 2 (6.7%) | - | - | 1 (2.9%) | 3 (2.4%) |
| Multiple | - | 1 (3.3%) | - | - | 1 (0.8%) |
| White | 28 (93.3%) | 29 (96.7%) | 30 (100.0%) | 34 (97.1%) | 121 (96.8%) |
| **Ethnicity** |  |  |  |  |  |
| Hispanic or Latino | 1 (3.3%) | 2 (6.7%) | - | - | 3 (2.4%) |
| Not Hispanic or Latino | 28 (93.3%) | 28 (93.3%) | 29 (96.7%) | 34 (97.1%) | 119 (95.2%) |
| Not Reported | 1 (3.3%) | - | - | 1 (2.9%) | 2 (1.6%) |
| Unknown | - | - | 1 (3.3%) | - | 1 (0.8%) |

Notes: Data are mean (SD) or n (%). LDR, low-dose duplicate recipient; HDR, high-dose duplicate recipient; HTR, high-dose triplicate recipient.

### 3.2 Vaccine Safety and Tolerability

As shown in Table 2, none of the treatment-emergent adverse events (TEAEs) resulted in the termination of participation in the trial. The majority of TEAEs were categorized as mild or moderate, and there was no observed escalation in either the dosage level or frequency of the vaccine. Primarily, a total of eight participants experienced TEAEs that were categorized as severe. Additionally, six participants reported TEAEs that were deemed significant, while one person chose to withdraw from the study due to TEAEs. Nevertheless, none of these events were directly associated with vaccinations.

**Table 2.** Overall Summary of TEAEs (Safety Population)

|  | **LDR**<br>**(N=30)** | **HDR**<br>**(N=30)** | **HTR**<br>**(N=30)** | **Pooled**<br>**Placebo**<br>**(N=35)** | **Overall**<br>**(N=125)** |
| --- | --- | --- | --- | --- | --- |
| **TEAEs** | 20 (66.7%) | 18 (60.0%) | 22 (73.3%) | 24 (68.6%) | 84 (67.2%) |
| Grade 1: Mild | 10 (33.3%) | 11 (36.7%) | 9 (30.0%) | 16 (45.7%) | 46 (36.8%) |
| Grade 2: Moderate | 8 (26.7%) | 6 (20.0%) | 10 (33.3%) | 6 (17.1%) | 30 (24.0%) |
| Grade 3: Severe | 2 (6.7%) | 1 (3.3%) | 3 (10.0%) | 2 (5.7%) | 8 (6.4%) |
| Grade 4: Life-threatening | - | - | - | - | - |
| **Serious TEAEs** | 2 (6.7%) | 1 (3.3%) | 1 (3.3%) | 2 (5.7%) | 6 (4.8%) |
| **TEAEs leading to withdrawal** | - | - | 1 (3.3%) | - | 1 (0.8%) |
| **TEAEs leading to discontinuation** | - | - | - | - | - |
| **Treatment-related TEAEs** | 6 (20.0%) | 8 (26.7%) | 7 (23.3%) | 7 (20.0%) | 28 (22.4%) |
| **Treatment-related TEAEs ≥ Grade 3** | - | - | - | - | - |
| **Treatment-related Serious TEAEs** | - | - | - | - | - |
| **Treatment-related TEAEs leading to withdrawal** | - | - | - | - | - |

Notes: LDR, low-dose duplicate recipient; HDR, high-dose duplicate recipient; HTR, high-dose triplicate recipient.

As shown in Figure 2, the occurrence of injection site bruising was the most common localized adverse event. The incidence rates were 3.3% (1 out of 30) in the low dose group (LDR), 3.3% (1 out of 30) in the high dose group (HDR), and 6.7% (2 out of 30) in the highest dose group (HTR). All adverse events occurring at the injection site were categorized as mild. The occurrence of headaches as a systematic adverse reaction was observed most frequently. Its prevalence was found to be 3.3% (1 out of 30) in the HDR group, 6.7% (2 out of 30) in the HTR group, and 2.9% (1 out of 35) in the pooled placebo group. The sole systematic adverse event categorized as moderate was fatigue in LDR, occurring at an incidence rate of 3.3% (1 out of 30).

**Figure 2.**
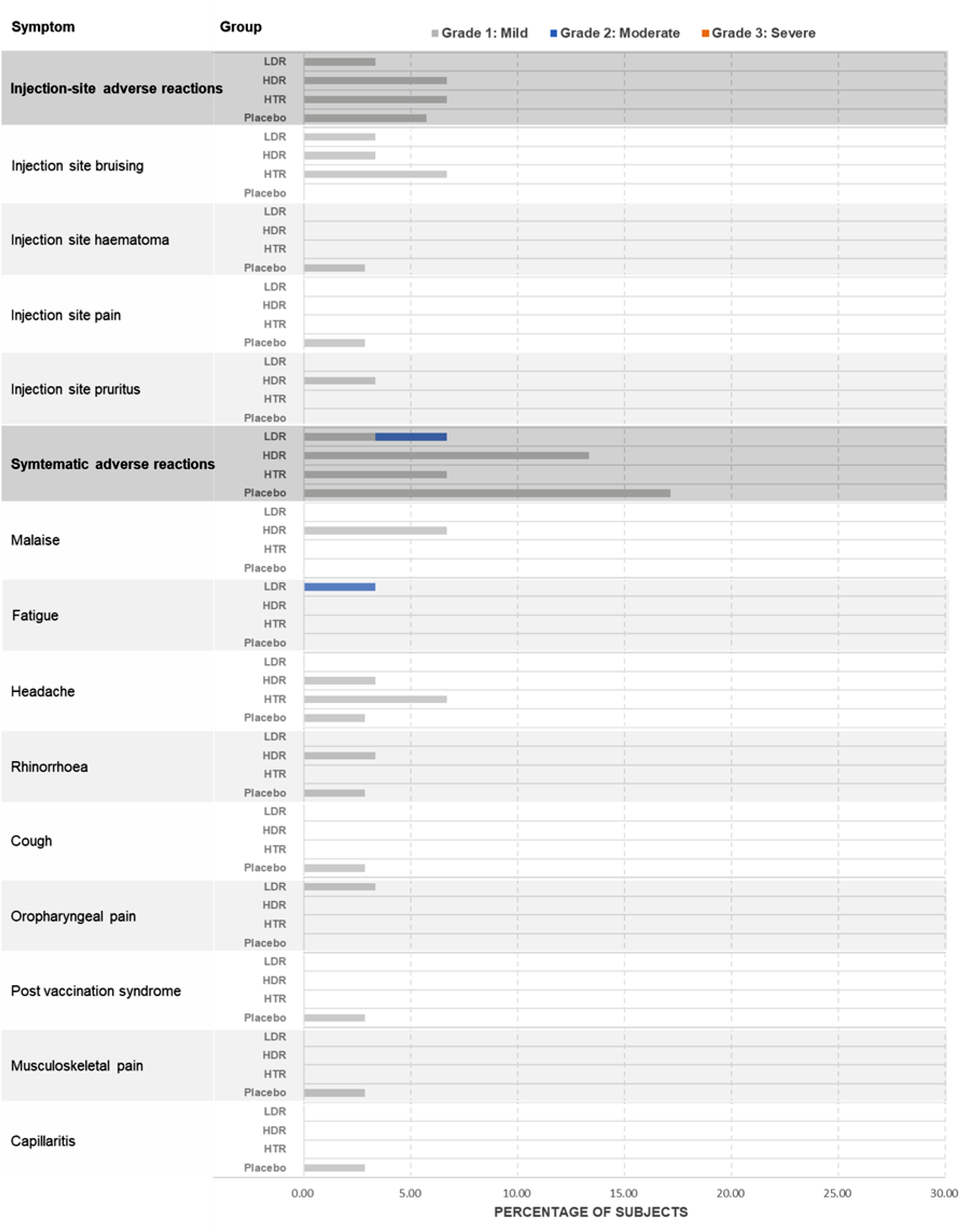
Summary of Treatment-related TEAEs for Maximum Severity (Safety Population) Treatment-related TEAEs in subjects who received BARS13 in low-dose duplicate, high-dose duplicate, or high-dose triplicate, or placebo, are depicted. Data for placebo have been pooled, as seen above. LDR stands for low-dose duplicate recipient; HDR stands for high-dose duplicate recipient; and HTR is for high-dose triplicate recipient.

### 3.3 Specific G Protein Binding Antibody Response

In the assessment of immunogenicity, it was shown that the geometric mean concentrations (GMC) of antibodies binding to the G protein increased after BARS13 vaccination in comparison to the baseline levels before immunization, across all groups. The initial geometric mean concentration (GMC) in the low-dose duplicate recipients’ group (Cohort 1, LDR) at Week 0 was determined to be 910.1 IU/mL, with a 95% confidence interval (CI) spanning from 675.1 to 2159.7. During the fourth week following the initial immunization, the GMC was recorded as 1051.3 IU/mL, with a 95% CI ranging from 836.5 to 2640.6. Subsequently, during the fourth week after the second immunization, the GMC was measured at 920.4 IU/mL, with a 95% CI of 438.7 to 2121.0 (Figure 3a). In the cohort consisting of recipients who received a high-dose duplicate (Cohort 2, HDR), the GMC was found to be 1513.0 IU/mL (95% CI: 1334.0-3503.2) four weeks after the initial immunization. Similarly, at the fourth week following the second immunization, the GMC was measured to be 1501.0 IU/mL (95% CI: 1049.2-3282.3). The observed values in this study exhibited a small increase compared to the pre-immunization baseline GMC of 1353.7 IU/mL (95% CI: 1224.6-2907.8), as depicted in Figure 3b. In Cohort 3, specifically the group of recipients who received high-dose thrice vaccines (referred to as HTR), the GMC at the beginning of the study (Week 0) was 881.0 IU/mL. The 95% CI for this GMC ranged from 794.5 IU/mL to 1473.4 IU/mL. During Week 4, the GMC was measured to be 1116.3 IU/mL, with a 95% CI ranging from 990.7 to 1772.5. At Week 8, the GMC increased to 1309.4 IU/mL, with a 95% CI of 1162.8 to 2041.5. Finally, at Week 12, the GMC further increased to 1359.6 IU/mL, with a 95% CI of 1197.9 to 2525.7. After each administration of BARS13, there was a notable and consistent rise in the levels of antibodies, as depicted in Figure 3c.

**Figure 3.**
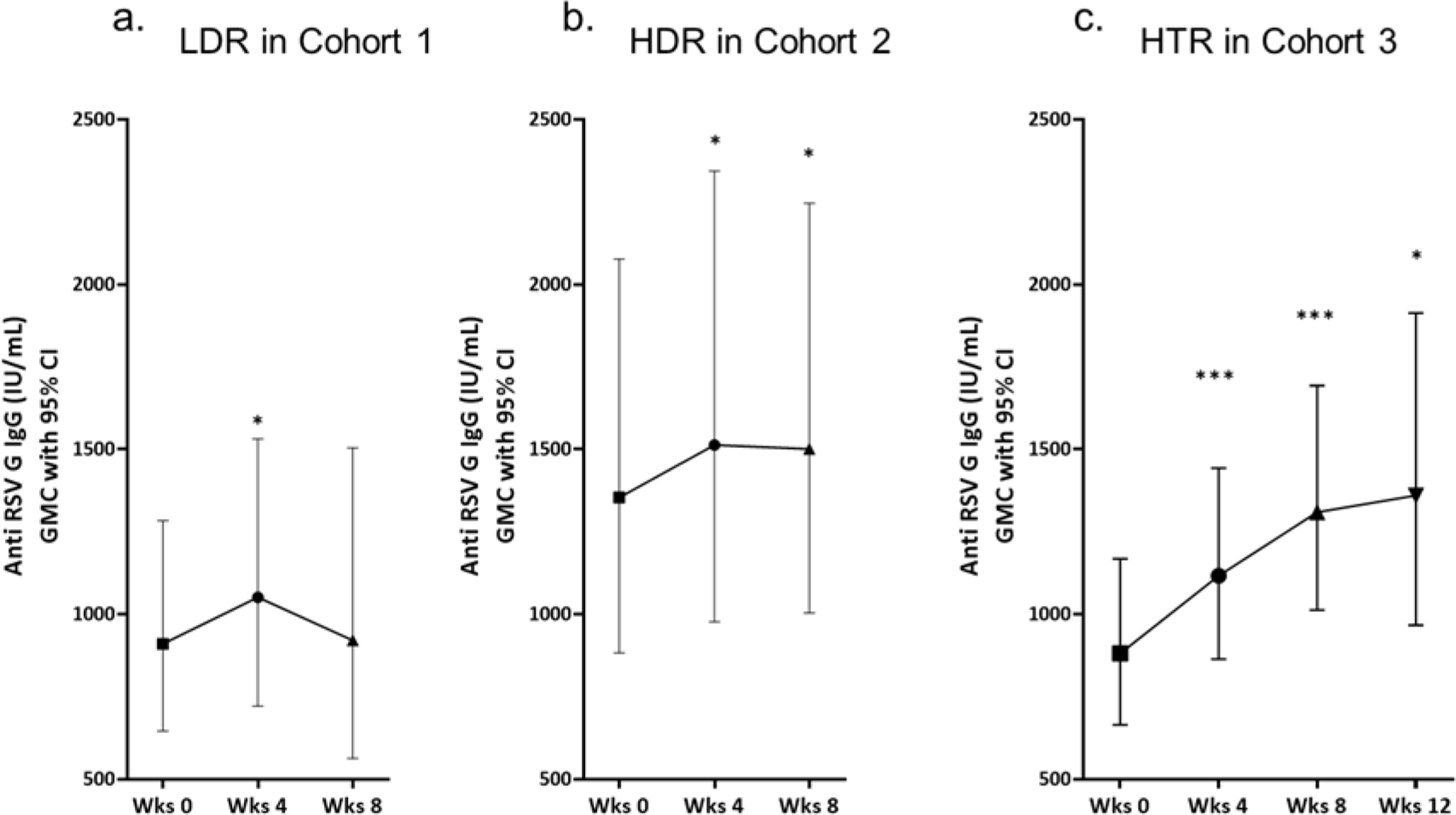
Analysis of Level of Anti-G Antibodies among Cohorts. Kinetics of vaccine-elicited anti-G responses shown as anti-G IgG GMCs with 95% CIs. Panels a and b show the humoral responses from 2-dose regimen of 10 µg BARS13 and 20 µg BARS13 respectively. Panel c shows the humoral responses of a triple-dose regimen consisting of 20 µg BARS13. The findings were presented using the GMC with a 95% confidence interval. The paired T-test was conducted, excluding estimated data from placebo individuals.

Participants who received a high dose regimen of twice 20 μg BARS13 (HDR) had slight higher antibody levels than those received a low dose regimen of twice 10 μg BARS13 (LDR). This finding suggests the presence of a dose-response correlation. Furthermore, with regard to the correlation between the total number of vaccinations and the immune response, it was observed that the levels of antibodies in HTR who received thrice of 20 μg BARS13 had a considerably superior performance compared to HDR in Cohort 2. This finding indicates that employing successive vaccination procedures, including increased dosages of BARS13, leads to enhanced production of anti-RSV antibodies.

The rate of responsiveness in individual participants has also been evaluated. The calculation and definition of the responsiveness rate involved determining the percentage of participants who exhibited an increase in antibodies relative to their pre-dose levels. For LDR in Cohort 1, the rate of responsiveness was found to be 84% at Week 4 which was 4 weeks after the initial dose and 83.3% at Week 8 which was 4 weeks after the second dose. The responsiveness rate for HDR in Cohort 2 was found to be 95.5% at Week 4 and 72.2% at Week 8. The responsiveness rate for HTR in Cohort 3 was seen to be 85.7% at Week 4, 85.2% at Week 8, and 79.2% at Week 12 which was 4 weeks after the third dose, as depicted in Table 3. The observed responsiveness rates provide an indication of the vaccine’s effectiveness in eliciting an immunological response, as demonstrated by the substantial proportion of individuals who produced antibodies against the specific antigen. The observed decrease in responsiveness rate among HDR in Cohort 2 following the second immunization could potentially suggest a marginal decline in immune response or could be attributed to other factors that have an influence. Nevertheless, it is worth noting that the responsiveness rates exhibit a consistently high level across all cohorts, indicating the vaccine is effective in stimulating an immunological response in the majority of participants. Additional studies are necessary to ascertain the most effective immunization schedule and evaluate the sustained impact of the immune response over an extended period.

**Table 3.**
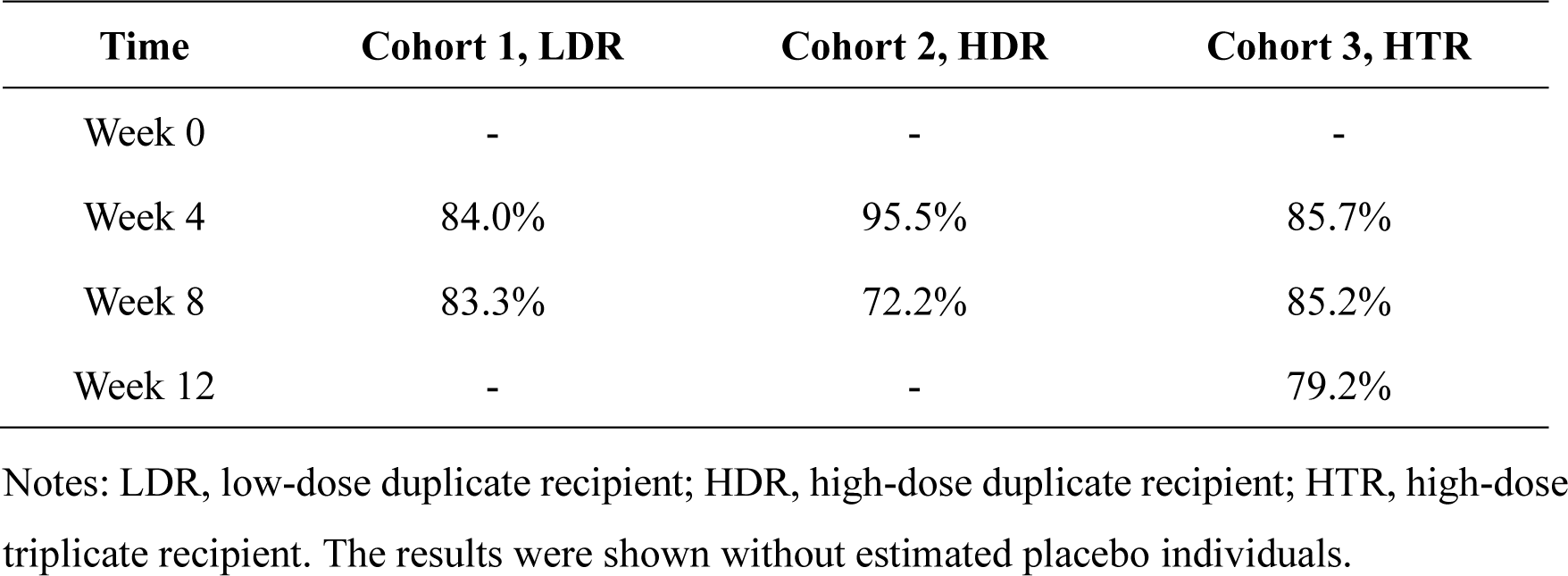
Analysis of Responsiveness to BARS13 Vaccinations among Cohorts.

According to the present analysis, it was shown that the treatment of BARS13 led to a significant and dose-dependent enhancement of humoral responses against RSV in all cohorts. In order to obtain a more comprehensive assessment of BARS13, we will proceed with the examination of humoral responses once the trial is no longer blinded.

## 4. DISCUSSION

The present study presents findings from the interim analysis of an ongoing Phase 2 clinical trial that aims to assess the safety, tolerability, and immunogenicity of BARS13, a recombinant G protein-based respiratory syncytial virus (RSV) vaccine combined with the immunomodulator CsA, in a population of older individuals. The study found that the BARS13 vaccine was administered to an older population with no serious adverse reactions. Additionally, the BARS13 vaccine induced a significant increase in IgG anti-RSV antibody levels in all cohorts, but higher doses and frequencies elicited stronger immune responses. Due to the limited sampling taken for the interim report, there was no duration of anti-RSV antibody assessment. Further analysis and evaluation are needed to fully understand the vaccine’s efficacy and long-term effects once the trial is officially closed and a full pack of data is available.

Among the 125 participants included in this interim analysis, no treatment-related serious TEAEs were reported in the safety population. No severe or life-threatening TEAEs were observed. A notable number of the TEAEs recorded in the study were classified as mild. A single instance of mild frontal fibrosing alopecia (an autoimmune condition) was reported by a recipient who received a high dose of the study medicine. This adverse event prompted the premature discontinuation of the therapy, although it was determined to be unrelated to the treatment. In general, the occurrence of mild and moderate TEAEs was comparable between those receiving the active vaccine and those receiving the placebo, and there was no observed correlation between TEAEs incidence and vaccine dosage. There was no observed correlation between the frequency of TEAEs and vaccine dosage level or frequency. Similarly, there was no observed rise in the frequency of treatment-related TEAEs with vaccine dose level or frequency. The majority of treatment-related TEAEs documented in this interim analysis report consisted of occurrences of headache and injection site bruising. In the older population, no clinically significant patterns of changes in the usage of concomitant drugs were observed among the various treatment groups.

The elderly population is frequently susceptible to respiratory syncytial virus (RSV) infections, which can pose a challenge to the efficacy of RSV vaccines. This is mostly attributed to the presence of pre-existing antibodies that neutralize the vaccine antigens, hence impeding the vaccination’s effectiveness. Developing an effective respiratory syncytial virus (RSV) vaccination for this particular age range presents a significant hurdle. Furthermore, the existence of pre-existing medical disorders and the concurrent use of medications may exacerbate the challenges associated with the effectiveness of the vaccine in older populations. Additionally, the immune system of older individuals tends to weaken over time, making it more difficult for vaccines to generate a strong and lasting immune response. This further emphasizes the need for careful evaluation and consideration of dosing escalation and frequency to ensure optimal effectiveness of an RSV vaccine in older populations. An analysis of the anti-RSV-G IgG antibody baseline revealed that subjects with a history of environmental exposure to RSV infections exhibited a significantly elevated baseline level of this antibody. To eliminate variables and inconsistencies among individuals, we employed the quantitative ELISA analysis to measure the concentration of anti-G IgG. These concentrations were reported in international units per microliter (IU/mL), along with 95% confidence intervals, for each treatment group during weeks 0, 4, 8, or 12.

The administration of two doses of low-dose BARS13 (10 µg) via injection produced significant anti-G IgG antibodies at Week 4 after vaccination. These antibodies decreased marginally until the present analysis, during which time more than 80% of the participants exhibited responsiveness. The cohorts were administered a high dosage of BARS13 (20 µg), resulting in the sustained production of antibodies at both Weeks 4 and 8. Furthermore, these antibodies exhibited an enhanced response towards BARS13. The administration of an additional dosage of BARS13 resulted in a notable increase in antibody production, with levels significantly above the initial baseline measurements at all observed timepoints. This demonstrated a dose-dependent increase pattern from the low-dose to high-dose cohorts, as evidenced by the rise from baseline to post-vaccination. As evidenced by the antibody levels in Cohort 3, which increased further after the second and third doses, the boost dose also played an important part in the antibody response.

A prior clinical study revealed that the importance of dosage was established in a dose-ranging investigation of extracted and purified F, G, and M antigens from RSV A viruses through intramuscular injection at doses of 100, 50, or 25 µg to the aged population. The antibody levels against RSV F and G proteins were found to be equal when administering doses of 25 and 50 µg of the vaccine. However, a considerably greater antibody response was achieved after administering a dose of 100 µg of the vaccine in comparison to the 25 and 50 µg doses. These findings suggest that a higher dosage of the vaccine may be necessary to elicit a stronger immune response in the elderly population. Further studies are needed to determine the optimal dosage for maximum protection against RSV. These results are significant as they indicate that the immune response to RSV can be enhanced by increasing the vaccine dosage. This is particularly important for the elderly population, as they are more susceptible to severe RSV infections. The findings warrant further investigation into the safety and efficacy of higher vaccine dosages and could potentially lead to the development of a more effective vaccine against RSV in older adults. Dosing selection is pivotal for G protein-based vaccines in the elderly population observed in this current study. The elderly population often has a weaker immune response to vaccines, making it necessary to administer higher dosages to achieve optimal protection against RSV. Additionally, our research suggests that dosing selection is critical in G protein-based vaccines for older adults, as it directly impacts the vaccine’s efficacy and effectiveness. These findings underscore the need for more research and development in this area to improve the immunity and health outcomes of the elderly population.

Overall, this phase II study in older adults showed different dose levels of the rRSV G protein investigational vaccine to be safe, well tolerated, and highly immunogenic and responsive in adults 60–80 years of age. Further research and development in this area is crucial in order to determine the optimal dosage and administration schedule of the RSV G protein investigational vaccine for maximum effectiveness in elderly individuals. Additionally, studying the long-term effects of the vaccine and its potential impact on reducing hospitalizations and mortality rates among older adults will provide valuable insights into improving their overall health outcomes. Ultimately, investing in the continued development and implementation of the RSV G protein vaccine has the potential to significantly enhance the immunity and well-being of the elderly population.

## 5. CONCLUSION

In summary, this phase 2 trial has demonstrated that BARS13, a recombinant G protein-based RSV vaccine, not only showed a well safety and tolerability profile across different dose groups in older population, but also, importantly, could induce a meaningful level of anti-G antibodies in a dose- and frequency-dependent mode. The continuous upward trend in antibody concentration up to the interim analysis is promising for the effectiveness of BARS13.

## Data Availability

All data produced in the present study are available upon reasonable request to the authors

## SUPPLEMENTARY MATERIALS

File S1. Study Vaccine Lots

## INSTITUTIONAL REVIEW BOARD STATEMENT

The study was conducted in accordance with the Declaration of Helsinki and approved by the Bellberry Human Research Ethics Committee (protocol code ADVA-BARS13-002 and approved on 01 February 2021) for the clinical trial. See more details at https://clinicaltrials.gov/study/NCT04681833?term=NCT04681833&rank=1.

## INFORMED CONSENT STATEMENT

Written informed consent has been obtained from the participant(s) to publish this paper.

## DATA AVAILABILITY STATEMENT

The data generated or analyzed during this study are included in this published article and its supplementary files.

## ACKNOWLEDGEMENTS

Advaccine is grateful to all trial participants, including the Principal Investigators (PI), Natasha Martin and Kristi Mclendon, the Contract Research Organization (CRO), Avance Clinical Pty Ltd., and the Central Laboratory, Agilex Biolabs Pty Ltd., for their important contributions to this research. We also recognize the outstanding and dedicated support of the clinical trial staff at two locations in Australia: CMAX Clinical Research Pty Ltd. in Adelaide and Nucleus Network Brisbane Clinic in Queensland. We are also grateful to our Advaccine team members who have steadfastly worked even during the COVID-19 pandemic.

