## Supplementary Files S1 for "A Phase 2 random, double-blind, placebo-controlled study of the safety and immunogenicity of a recombinant G protein-based respiratory syncytial virus vaccine in healthy older adults"

**Supplementary Materials**

**File S1. Study Vaccine Lots**

The following lot numbers of the study vaccines and diluents were used: Lyophilized rRSV-G protein (R202101); CsA diluent (C202002); Placebo for rRSV-G protein (R202002K); Placebo for CsA (C2012001K). Lyophilized rRSV-G protein and placebo for lyophilised powder IPs were stored at < -15 to -25°C. Diluents were stored at 2 to 8°C. Prepared syringes containing reconstituted vaccine were used within 1 hour of preparation at ambient room temperature or stored at 2-8°C for a maximum of 4 hours prior to use.
